# Eco-emotions and suicidal ideation and behaviour: A systematic literature review

**DOI:** 10.1101/2023.12.15.23300020

**Authors:** Kairi Kõlves, Damian Shaw-Williams, Sadhvi Krishnamoorthy, Sharna Mathieu, Linda R. Zhong, Aarthi Ganapathy, Jacinta Hawgood, Caroline Donovan, Susan H Spence, Lennart Reifels

## Abstract

**Introduction:** Although there have been assumptions about the link between eco-emotions and suicidality, there is currently no systematic evidence. Therefore, the aim of this review is to systematically examine empirical literature analysing the link between eco-emotions and suicidal ideation and behaviours, both fatal and non-fatal.

**Methods:** The review protocol was pre-registered in PROSPERO [CRD42022352379] and the PRISMA checklist was followed. Searches were conducted in six electronic databases (Scopus (Elsevier), Medline (PubMed), Web of Science, ProQuest, CINAHL and Embase) for peer-reviewed English language literature published from 1 January 2000 until 16 September 2023. Screening of titles, abstracts and full texts was conducted independently by two reviewers and any discrepancies were resolved in consultation with a senior researcher. The Johanna Briggs Institute’s quality appraisal tools were used for quality assessment.

**Results:** Search results revealed 559 records. After removal of duplicates, 424 articles were screened. After title and abstract screening, 28 articles were included for full text screening. Only one paper satisfied the inclusion criteria. A cross-sectional online knowledge, attitudes, and practice (KAP) survey among practicing mental health professionals across the State of Minnesota analysed the impact of climate change on their work and clients. Although the study did not specifically focus on suicidality, it reported that 22% of practicing mental health professionals had seen evidence of suicidal ideation or attempts in their clients as an outcome of climate change.

**Conclusion:** There is a lack of research on the association between eco-emotions and suicidality. Considering advancing climate change research, our review emphasizes an urgent need to conduct high level research to analyse the association between eco-emotions and suicidality and consider its potential implications.

**What is already known on this topic?:** There’s a recognized and growing concern about the impact of climate change on mental health, particularly the emergence of eco-emotions such as eco-anxiety, eco-grief, and solastalgia. While studies have linked higher temperatures and natural disasters to suicidality, research specifically examining the association between eco-emotions and suicidal ideation and behaviours is lacking.

**What this study adds?:** This systematic literature review revealed a stark scarcity of research connecting eco-emotions with suicidal ideation and behaviours. Only one paper met the inclusion criteria of the review. This scarcity underscores the urgent need for more comprehensive studies exploring the potential connection between eco-emotions and suicidality at an individual level.

**How this study might affect research, practice, or policy?:** The limited findings from this review highlight the pressing need for more in-depth research to determine and understand the association between eco-emotions and suicidal ideation and behaviours. It emphasizes the necessity for interdisciplinary research for a multi-faceted approach addressing mental health impacts of climate change and its underlying causes, calling for informed engagement, mental health support, and policy actions to mitigate eco-grief and anxiety related to climate change impacts.

## Introduction

Climate change is a complex and multifaceted issue that poses significant threats to ecosystems, human health, and the global economy.^1^ As the impacts of climate change become increasingly apparent, there is growing concern about their effect on mental health.^2-4^ New concepts are emerging in the field of environmental psychology, including eco-emotions, which describe emotional responses to perceived threats and losses associated with climate change. The most described eco-emotions in the literature include eco-anxiety, eco-grief, and solastalgia.^5^

Eco-anxiety is a term used to describe the psychological distress caused by climate change and environmental degradation.^6^ It is an emotional response that may encompass worry, guilt, sadness, and hopelessness in response to the current climate crisis. Eco-anxiety can also lead to proenvironmental behaviours.^7^ As defined by the American Psychological Association, eco-anxiety is ‘a chronic fear of environmental doom,^8^ ranging from mild stress to clinical disorders like anxiety and depression.^6^ Nevertheless, further research is needed for the clarity of the definition.^9^

Eco-grief refers to the sense of loss that arises from experiencing or learning about environmental destruction or climate change.^10^ It is a form of grief related to concerns about the state of the world’s environment, including the loss of ecosystems, landscapes, species, and ways of life.^11^ Ecological grief can be considered a form of “disenfranchised grief” or a grief that is not publicly or openly acknowledged.^10^ It can be characterized as falling into three main areas: grief associated with the loss of species and biodiversity, grief associated with the loss of ecosystems and landscapes, and grief associated with the loss of cultural heritage and ways of life.^12^ Ecological grief can pose a mental health risk but can also be a functional response to environmental change.^11.12^

Solastalgia, a term coined by Albrecht,^13^ is often described as a form of ecological grief. While solastalgia and eco-grief share some similarities, they may be associated with different types of environmental change and emotional responses. Solastalgia is more closely tied to a sense of place and specific location, while eco-grief may be more broadly associated with the loss of biodiversity or ecosystem health. As defined by Clayton et al.,^8^ solastalgia is ‘the lived experience of negatively perceived change to a home environment’ (p. 27).

The prevalence of eco-emotions is likely to increase as the impacts of climate change become more pronounced and widespread, potentially having significant implications for mental health and, in extreme cases, leading to suicidal ideation and behaviours.^5,6,14^ A recent Lancet seminar by Knipe et al.^15^ highlighted climate change as a key global risk factor of increasing importance for suicide and self-harm. What is the current evidence? Research shows that higher temperatures affect mental well-being, furthermore, higher suicide rates are associated with an increase in temperatures and drought in both developed and developing countries.^4,16-19^ There have also been systematic literature reviews on the potential impact of natural disasters (which are likely to increase in frequency and severity as a result of climate change) on suicidal ideation and behaviours, which have shown inconclusive results (e.g., Kolves et al.,^20^ currently updated CRD42020216722). However, while there have been assumptions about the link between eco-emotions and suicidality, there have been no systematic reviews on this topic. Therefore, the aim of this review is to systematically examine empirical literature that explores the link between eco-emotions and suicidal ideation and behaviours, both fatal and non-fatal.

## Methods

The presentation of this systematic review adheres to the Preferred Reporting Items for Systematic Reviews and Meta-Analyses statement^21^ (PRISMA checklist is provided in Supplementary Table 1). No funding agency played a role in study design, data collection, data analysis, data interpretation, or report writing. The review protocol was pre-registered in PROSPERO [CRD42022352379].

### Inclusion/exclusion criteria

We included all study designs that presented primary empirical (both quantitative and qualitative) analyses of eco-anxiety, eco-grief, and other related climate emotions in connection with suicidal ideation and behaviours (both fatal and non-fatal). Studies addressing the potential impact of specific events such as natural disasters (e.g., floods, landslides, droughts) and man-made disasters (e.g., industrial accidents, acts of terrorism, war, or other incidents of mass violence/casualty) were excluded unless they measured also eco-emotions. While recognizing that such events can contribute to significant ecological harm and may be associated with climate change, it is essential to note that individuals can experience eco-anxiety and eco-guilt without direct traumatic exposure to extreme disaster events. Furthermore, disasters may directly contribute to suicidal ideation and behaviour (for a comprehensive review, see Kolves et al.^20^). We also excluded studies that did not present suicidal ideation or behaviours as an outcome or associated factor.

### Search strategy and selection criteria

A systematic search of six electronic databases was conducted for peer-reviewed English language literature published from 1 January 2000 until 16 September 2023. Databases included Scopus (Elsevier), Medline (PubMed), Web of Science, Proquest, CINAHL and Embase. Search terms included the following:

suicid* OR self-harm OR self-injur* (All fields)

AND

“climate anxiety” OR “eco-anxiety” OR “ecological anxiety” OR “ecological grief” OR “eco-grief” OR “climate worr*” OR “climate fear*” OR solastalgia OR “eco-guilt” OR “environment* despair” OR “eco-depression” OR “ecological depression” (Title, Abstract, Keywords).

### Screening

Descriptive data were exported from the databases into a pre-extraction file to record authors, title, year of publication, journal/volume/issue, and article abstract. Duplicate records were removed. Independent reviewers (SK, LZ, AG) screened the titles and abstracts against the eligibility criteria. Any discrepancies were resolved through discussions with the project lead (KK). Following this, the remaining full-text articles were independently reviewed by reviewers (SK, LZ) to determine final eligibility. Any discrepancies were resolved through discussions with the project lead (KK), who also reviewed the respective papers. Reference lists of the included records were manually screened by KK to identify further eligible studies.

### Data extraction and analysis

Data extraction was carried out by one reviewer (KK) and cross-checked by another reviewer (DSW). The following information was extracted from the included articles: Authors and Date of publication, Data Collection Time Period, Population, Study Design, Measure of Eco-emotion (or related construct[s]), Measure of suicidal ideation and behaviour, and Main finding(s), such as effect measures or qualitative themes. Additionally, data on the country, sample size, funding source, and main limitations were extracted. The information was synthesized into narrative summaries as meta-analysis was not feasible due to limited results.

### Risk of bias (quality) assessment

Study level quality was appraised using the suite of quality appraisal tools developed by the Johanna Briggs Institute.^22^ Two independent reviewers (KK & SK) appraised each study. Studies were not excluded based on quality appraisal.

## Results

Search results from six databases revealed 559 records (see details in Figure 1). We removed 135 duplicates, leaving 424 articles for screening. Based on title/abstract screening, 396 records were excluded and the full texts of 28 articles were read. Following this, only one paper satisfied the inclusion criteria.

**Figure 1.**
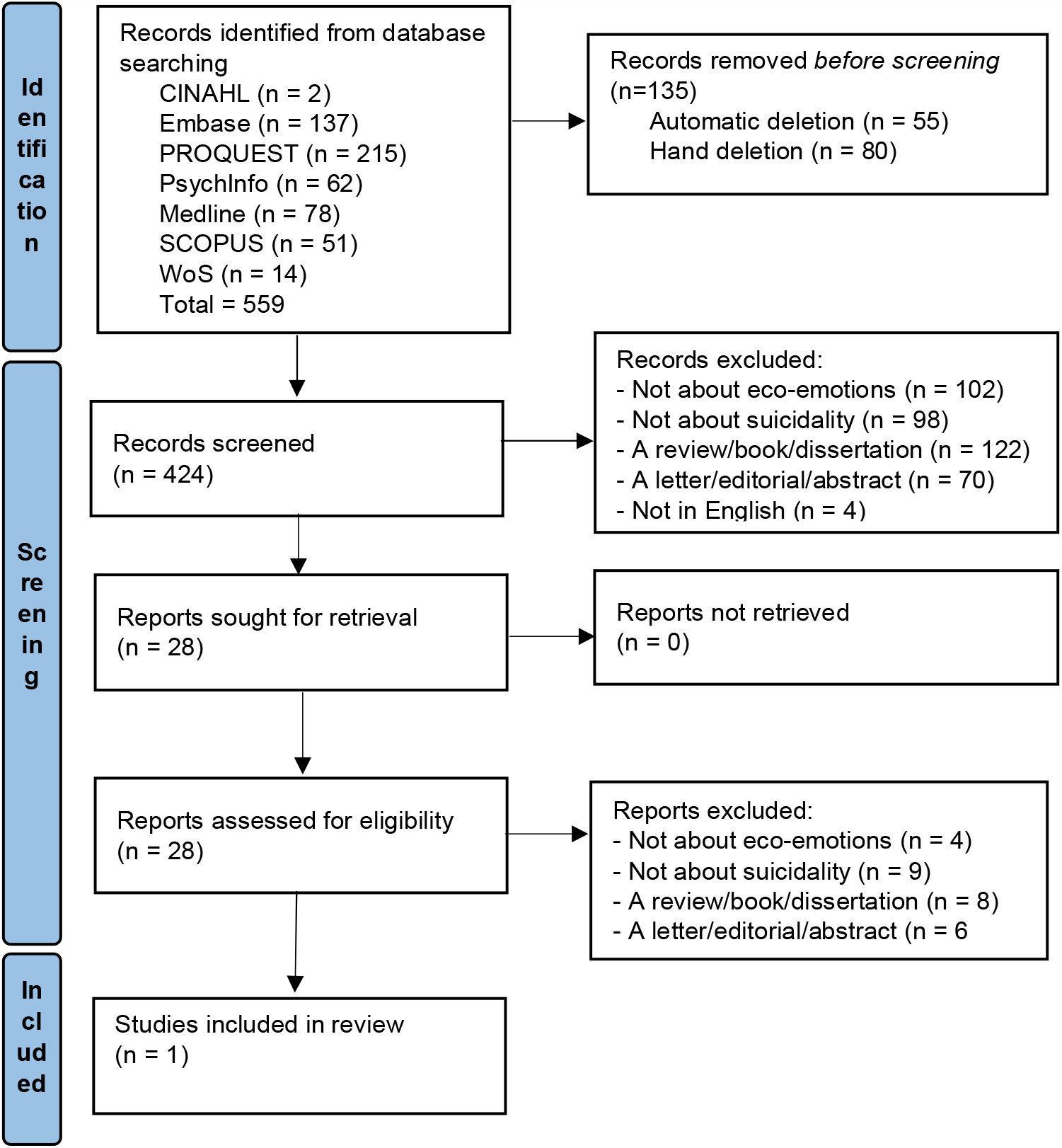
Study selection flow-diagram based on PRISMA guidelines.

- Figure 1 in here please

The paper by Hoppe et al.^23^ focused on the challenges faced by licensed practicing mental health professionals (MHPs; Supplementary Table 2). The study used a cross-sectional online knowledge, attitudes, and practice (KAP) survey among practicing MHPs across the State of Minnesota. The survey revealed that out of 517 practicing MHPs, 81.6% agreed that climate change is an important problem impacting mental health, with many already observing these impacts. While the study did not specifically focus on suicidality, their list of mental health outcomes due to their clients’ climate change-related concerns included suicidal ideation or attempts. Over one-fifth (22%) of practicing MHPs reported that they had seen evidence of suicidal ideation or attempts in their clients as an outcome of climate change.^23^ It is important to note that the publication lacks important details about the design and conduct of the study, and there are some limitations. Although sample size calculations were provided, the information presented suggests that an online survey was shared with members of professional organizations, and random sampling was not conducted, which would be required to determine the scope of the problem. Furthermore, the response rate was not provided, and the sample description lacked information about gender and age. Data analysis was rather limited, presenting frequencies. Although the questionnaire was developed by a group of experts in climate and (mental) health, information about suicidal ideation and attempts was entirely based on practitioners’ perceptions.

## Discussion

The impacts of climate change present a growing emotional toll, resulting eco-emotions may lead to mental health problems and suicidality. This is the first systematic literature review to analyse the association between eco-emotions such as eco-anxiety, eco-grief and solastalgia with suicidal ideation and behaviours. We identified 424 unique papers with searches in six databases and only one paper could be included. The only study eligible for inclusion in the current review was by Hoppe et al.^23^ which focused on knowledge, attitudes, and practice in relation to climate change among MHPs in Minnesota, USA. Among other items, their questionnaire included a section about

MHPs’ experience regarding climate-related impacts on clients’ mental health including suicidality. Based on their perception, over one-fifth of MHPs had clients experiencing suicidal ideation or attempts as an outcome of climate change. While being limited to English-language publications since 2000, we did not identify any studies specifically analysing the association between eco-emotions and suicidality at an individual level as opposed to the views of MHP’s of their clients. This glaring gap in the research landscape underscores the urgent need for further studies to explore the potential connection between eco-emotions and suicidality.

The findings by Hoppe et al.^23^ should be interpreted within the context of the scarcity of similar studies. Nevertheless, as the authors point out, while climate change may be observed in Minnesota, the prevalence of mental health outcomes might be even higher in regions with greater exposure to natural disasters, including floods and droughts. It is crucial to explore various pathways to eco-emotions and, subsequently, to suicidal ideation and behaviour. Heightened eco-anxiety and concern for climate change could potentially link to anxiety proneness and generalized anxiety disorder. However, Pihkala^24^ notes that while traits like anxiety sensitivity contribute to eco-anxiety, they do not fully explain it. There is a need for deeper investigations into the multifaceted factors involved. For example, certain life situations, such as exposure to natural disasters, and specific occupations like farming, elevate susceptibility to eco-anxiety, regardless of individual personality traits. There is still much to understand about the connections between eco-emotions and suicidal ideation and behaviours. Questions arise: Is there a definite link between eco-emotions and suicidality? Could this relationship be causal? Are mental health issues mediating this connection? And could age and gender play a moderating role? Addressing these research queries is crucial, while acknowledging the challenges inherent in conducting high-quality suicide research, encompassing definitional complexities and the statistical rarity of suicide.^25^

Another important aspect for future research is measuring eco-emotions. Early scales in the field were focussed on environmental or climate distress (e.g., Environmental Distress Scale^26^). In recent years, a few scales have been developed specifically to measure eco-emotions. A comprehensive search by Cianconi et al.^5^ identified ten validated psychometric tests for assessing eco-emotions and psychoterratic syndromes; measuring specifically eco-anxiety, eco-grief and solastalgia (e.g., The Hogg Eco-Anxiety Scale^26^) Nevertheless, further testing, validation and use of those scales in surveys, and ideally in cohort studies including also suicidal ideation and behaviour, is encouraged.

Despite lack of studies specific to eco-emotions and suicidal ideation and behaviour, we should consider potential implications. As Hoppe et al.^23^ noted, only a small proportion of MHPs were familiar with resources for assessing and dealing with climate related emotions, and even fewer had utilized relevant tools with their clients. There is need for interdisciplinary research and collaborative practices aimed at designing and implementing assessment, intervention, and evaluation tools that can effectively address the impacts of climate change on individuals seeking help. Furthermore, there is need to develop mental health and suicide strategies to enhance community resilience in the face of climate-related stressors.^4^

Some potential implications include increasing awareness and understanding of the psychological impacts of climate change, and providing individuals with the tools and resources they need to cope with their feelings of anxiety and distress.^8^ This may include providing access to mental health services, as well as promoting mindfulness, meditation, and other stress-reducing activities. Ecotherapy has been recommended as a promising intervention.^23^ Ecotherapy is based on the idea that people are connected to and impacted by the natural environment; it involves experiencing nature to remediate mental health symptoms and boost overall well-being.^28^ The practice of ecotherapy may involve varying amounts of physical activity, depending on the type of program, and can include activities that focus on working in nature, experiencing nature, and spending time with others in nature. Some of the more common ecotherapy activities include nature meditation, gardening, hiking, and animal-assisted therapy. Nevertheless, more research about its impact and cost-effectiveness is required.^29^

At the policy level, the feelings of powerlessness and of being overwhelmed by the scale of the issue can form the basis of a call to action, and some measure of agency can be regained by being informed and involved in the policy process.^4^ A recent Australian study showed that eco-anger is a key emotional driver of engagement with climate action, while eco-anxiety and eco-depression were likely to be linked with disengagement and mental health problems.^30^ Nevertheless, Lester^31^ called for leading international and national suicide prevention organisations to urge governments to promote policies for mitigating climate change. Overall, a comprehensive policy response to eco-grief, eco-anxiety, and suicidality related to the impacts of climate change would require a multifaceted approach that addresses both the mental health impacts of climate change and the underlying causes of this issue.^12^

In conclusion, our systematic literature review explored the link between eco-emotions, such as eco-anxiety, eco-grief, and solastalgia, and suicidal ideation and behaviours. The study identified a notable scarcity of research in this area, with only one paper meeting the inclusion criteria. This highlights an urgent need to understand and ultimately address the potential connection between eco-emotions and suicidality, especially as the impacts of climate change escalate.

## Data Availability

All data produced in the present work are contained in the manuscript

